# Recovery Ratios Reliably Anticipate COVID-19 Pandemic Progression

**DOI:** 10.1101/2020.04.09.20059824

**Authors:** Dan V. Nicolau, Alexander Hasson, Mona Bafadhel

## Abstract

The COVID-19 pandemic is placing unprecedented demands on healthcare systems worldwide and exacting a massive humanitarian toll. This makes the development of accurate predictive models imperative, not just for understanding the course of the pandemic but more importantly for gaining insight into the efficacy of public health measures and planning accordingly. Epidemiological models are forced to make assumptions about many unknowns and therefore can be unreliable. Here, taking an empirical approach, we report a 20-30 day lag between the peak of confirmed to recovered cases and the peak of daily deaths in each country, independent of the epoch of that country in its pandemic cycle. This analysis is expected to be largely independent of the proportion of the population being tested and therefore should aid in planning around the timing and easing of public health measures. Our data also suggests broad predictions for the number of fatalities, generally somewhat lower than most other models. Finally, our model suggests that the world as a whole is shortly to enter a recovery phase, at least as far as the first pandemic wave is concerned.

## Introduction

The COVID-19 pandemic, at the time of writing, has infected at least 1.5millon people, and caused >100,000 deaths. The development of models to understand and predict the course of COVID-19 is imperative, in order to gain insight into the efficacy of public health measures aimed at containing its spread. Current models are either epidemiological^1,2^ or based on reported infection data^3^. These both have limitations: epidemiological models are forced to make assumptions about many unknowns thus varying wildly in their predictions, whilst reported data are retrospective and thus not predictive. It would be advantageous to have models that are directly data-driven and thus not forced to make assumptions, while retaining the predictive aspect of epidemiological models.

## Methods

To this end, we performed an empirical analysis of recovery trends around COVID-19 in individual countries and globally. We obtained our data from the Johns Hopkins Coronavirus Tracker^5^. We considered the ratio of known documented cases to recovered cases (C:R), in addition to the number of daily reported deaths (D). Since mortality is low (perhaps in the order of 1%), C:R eventually tends towards approximately 1 in the long term, indicating pandemic resolution.

We also considered the number of daily deaths recorded in each country, averaged over a 72-hour window (in order to reduce reporting stochasticity in the data). In those countries where this peak (as far as a first pandemic wave is concerned) has passed, we are then theoretically able to broadly predict the final total fatalities by projecting the area under the D curve.

## Results

The results of our analysis for all countries is available at the website covidwave.org. Figure 1 below shows C:R (blue) and D (red) for a selection of countries, chosen to be at various stages of the pandemic cycle, from different continents and with differing levels of healthcare access for the public. We are able to make several broad observations on the basis of these data.

**Figure 1.**
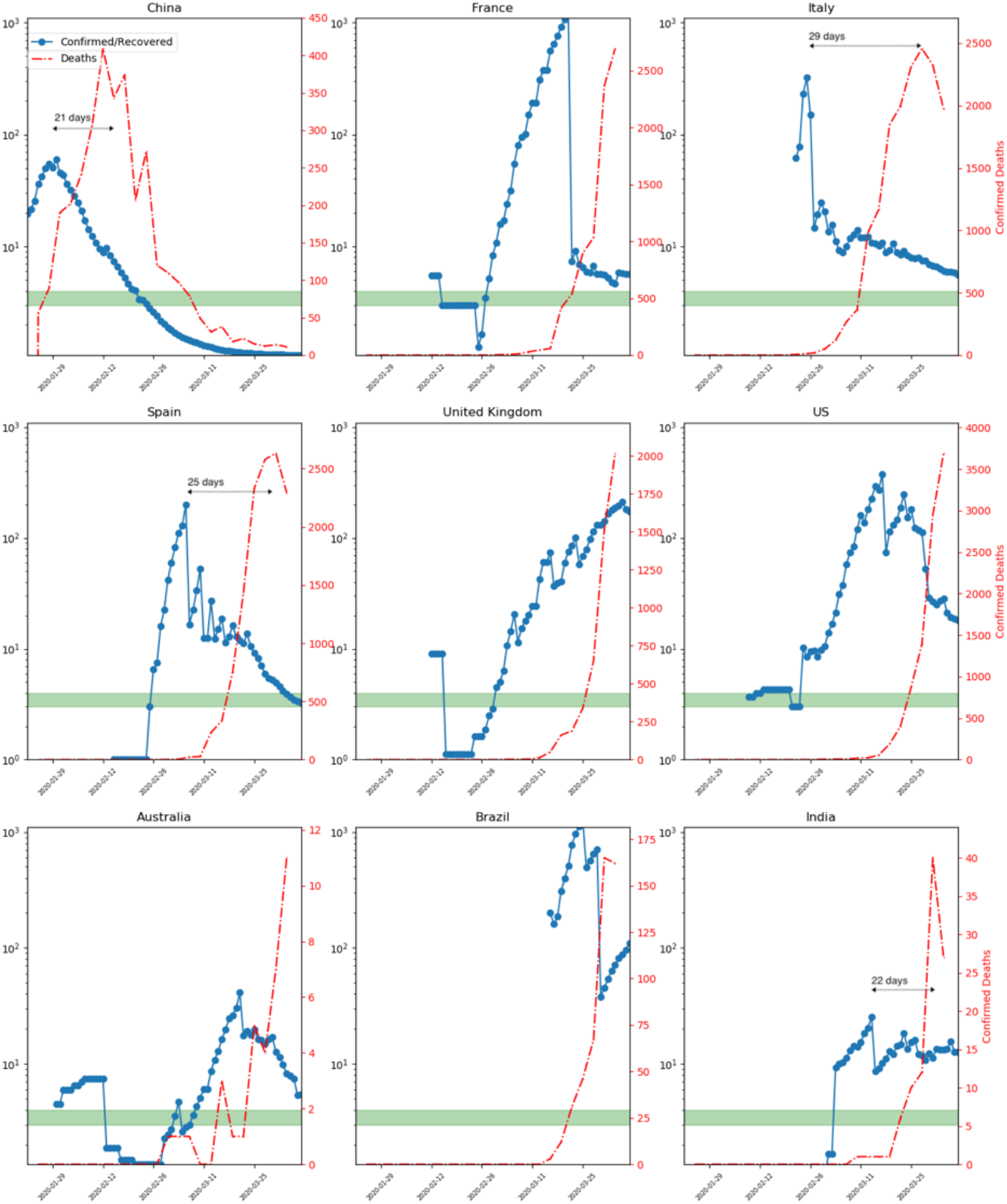
Blue plot: ratio of recorded to recovered cases (C:R). Red plot: daily deaths (72 hour running average). C:R tends towards 1 over time and its entering the green zone (2.0-5.0) gives an indication of when the first wave is expected to resolve. Note C:R anticipates the peak of daily deaths by 20-30 days in all cases where this peak has occurred. The area under the red curve predicts final death toll. Full data for all countries at covidwave.org.

**Firstly**, virtually all countries follow the same pattern, with the R:C curve climbing to a peak and then inverting once it passes a level in the mid-hundreds, but at different epochs. Our data suggest that once the C:R curve enters the green band (C:R between 2-5, the start of the resolution phase), on its way towards a limit of ∼1, there is an expectation that the (first) pandemic wave has passed. Conversely, a rising C:R portends a period of relative crisis; the UK and Brazil are in this phase. Our model suggests that Italy, Spain and Australia will enter their period of resolution by the second half of April. The USA represents an intermediate case, at an earlier stage in its relative recovery.

**Secondly**, we observe that the peak C:R anticipates the peak of daily reported deaths by ∼25 ± 5 days, with remarkable consistency between countries (Fig 1, arrows indicating the lag between the peaks of C:R and D). These data broadly predict a (first-wave) death toll of ∼25,000 and ∼50,000 for the UK and US, respectively and also that the first COVID-19 death-free days will occur in July for both.

**Finally**, these data indicate that SARS-CoV-2 spread quietly and globally for a significant period of time, mostly unnoticed. In this context, it is clear that the public health response of most authorities was substantially delayed. There are a few notable exceptions, including Singapore, South Korea and New Zealand and this is visible in the respective plots for these countries through the relative absence of a clear C:R peak, replaced by an early and prolonged plateau in this metric.

## Discussion

This analysis confirms that the COVID-19 pandemic behaves in similar ways across countries and can demonstrate a response of an individual national authority on recovered cases and deaths. These predictions also take into account current nationwide practices to halt spread and can lead to decisions to change how this is done and timed, including relaxing or tightening restrictions^4^.

With respect to the world as a whole, the respective plot (top left panel, covidwave.org) is difficult to interpret because it combines data from many countries with very different populations and at different epochs within the pandemic cycle. Nonetheless, one can identify a roughly 23 day lag between the first C:R peak (mainly mainland Chinese patients) and the first D peak (again mainly in Chinese patients), in keeping with our observations above. The second wave, now a global one, appears to be peaking approximately at the time of writing (mid-April, 2020) while there concomitantly appears to be a reduction in the number of deaths per day globally. If sustained, this early relative recovery, together with the earlier peak, would indicate a final global number of COVID-19 fatalities in the order of 250,000.

These data and projections are, naturally, subject to limitations. The most important of these relate to whether there will be a large rebound in cases nationally and globally once restrictions on movement and gatherings are relaxed. Our analysis cannot predict this aspect. There is also uncertainty around whether, independent of rebounds connected to the easing of public health measures, there would be a resurgence in COVID-19 cases in the Northern Hemisphere autumn, as is often the case with seasonal influenza.

## Data Availability

All data freely available at covidwave.org.

https://www.covidwave.org

